# In situ pressure modulation measurements by video capillaroscopy reveal capillary stiffening and reduced reactivity with age

**DOI:** 10.1101/2024.11.01.24316613

**Authors:** Marcus L. Forst, Gabriela Rincon, Juliette H. Levy, David N Cornfield, Stephen R Quake

## Abstract

Understanding tissue degradation with age and disease requires a clear picture of capillary function, but the small size of capillaries makes them difficult to model, perturb, and image. To measure capillary reactivity and stiffness, we recorded videos of nailbed capillaries while applying increasing external pressure to the finger. Combining the velocities at each pressure into participant velocity distributions showed that participants aged fifty and over (n=19) had a significantly higher median velocity than participants under fifty (n=16), whereas the effects of sex and blood pressure were insignificant. Notably, the effects of age were independent of blood pressure, suggesting that this method probes a different aspect of vascular health. Calculating dissimilarity using the earth-mover’s distance between individual and population velocity distributions classified age groups above and below fifty with an AUC of 0.79*±*0.07. These findings show that pressure-applying video capillaroscopy can measure the effects of capillary stiffening with age and interrogate how individual capillaries respond to the slowing and stopping of blood flow—suggesting that this method could be used to diagnose and evaluate vascular diseases in the future.

## 1 Introduction

The cardiovascular system operates as a feedback loop, adjusting blood flow based on body position and activity level. As people age, this system begins to break down as vessels become more stiff [1–4], causing downstream degenerative neurological diseases [5, 6], hypertension [7], heart attacks, and strokes [8]. Current vascular health assessments ranging from non-invasive blood pressure measurements to invasive cardiac catheterization [9–17] evaluate heart, artery, and vein function, but overlook capillaries because of their difficulty to image and model. Capillary images have long been used to classify Raynaud’s Syndrome [18]–a common disorder of poor capillary circulation–but their use has been confined to morphological dermatology assessment. But, recent studies in age-related diseases–such as systemic sclerosis[19, 20], diabetes[21, 22], chronic kidney disease [23, 24], and hypertension [25]–have found that capillary damage and dysfunction precede systemic symptoms. This suggests that capillary assessments may enable early detection and intervention for vascular diseases. Recent advances in camera and computer vision technology have enabled high spatiotemporal-resolution videos of capillary blood flow, known as ’video capillaroscopy.’ This technique shows promise in detecting neutropenia in nailbed capillaries[26], classifying white blood cell types in ventral tongue capillaries[27], identifying rolling inflammatory white blood cells in human oral mucosa capillaries [28], and measuring pulse-wave propagation in retinal capillaries[29]. This work seeks to use video capillaroscopy to extend the extensive research on systemic vascular aging; evaluating healthy adults of many ages and identifying changes in stiffness and vascular reactivity. We present ’pressure-applying video capillaroscopy,’ a non-invasive method to measure the response of nailbed capillaries to external pressure. By squeezing the finger with increasing pressure against a coverslip using an inflatable ’finger lock,’ we progressively slow down and eventually stop blood flow. Using video-analysis software, we quantify when blood flow slows, stops, and returns in a diverse population of healthy adults (n=35, 17 male, 18 female) and identify changes in capillary function and reactivity with age. Our results establish the groundwork for a non-invasive diagnostic test with early detection potential for vascular conditions and diseases.

## 2 Results

### 2.1 Procedure

We enrolled 35 healthy participants across ages (16 under fifty and 19 above fifty) and sex (17 male and 18 female) (Table 1). Participants carried out their normal routines before their visit. At the beginning of the visit, participants rinsed their hands under 35.5C water for one minute to normalize hand temperature for different seasons and people. To assess the response of capillaries to compressive force, we recorded videos of nailbed capillary blood flow using a custom-built capillary microscope (Fig. 1 a-b; Methods) which applies external pressure to the finger using an inflatable nitrile glove. We increased pressure every 10.49 seconds from 0.2 psi to 1.2 psi in 0.2 psi steps– gradually slowing blood flow–before reducing the pressure from 1.2 psi to 0.2 psi in the same manner. We then measured the participant’s blood pressure and repeated the procedure imaging a different set of capillaries in the same or a different finger (Methods).

**Table 1.** Participant Demographics and Characteristics

| Group | Total Participants (Mean Age ± SD) | BP < 120 sys | BP ≥ 120 sys |
| --- | --- | --- | --- |
| Total | 35 (46.1 ± 21.4) | 20 (57%) | 15 (43%) |
| Under Fifty | 16 (25.2 ± 5.8) | 10 (62%) | 6 (38%) |
| Above Fifty | 19 (63.8 ± 10.9) | 10 (53%) | 9 (47%) |
| Male Participants | 17 (45.4 ± 24.1) | 8 (47%) | 9 (53%) |
| Female Participants | 18 (46.9 ± 19.2) | 12 (67%) | 6 (33%) |

**Fig. 1.**
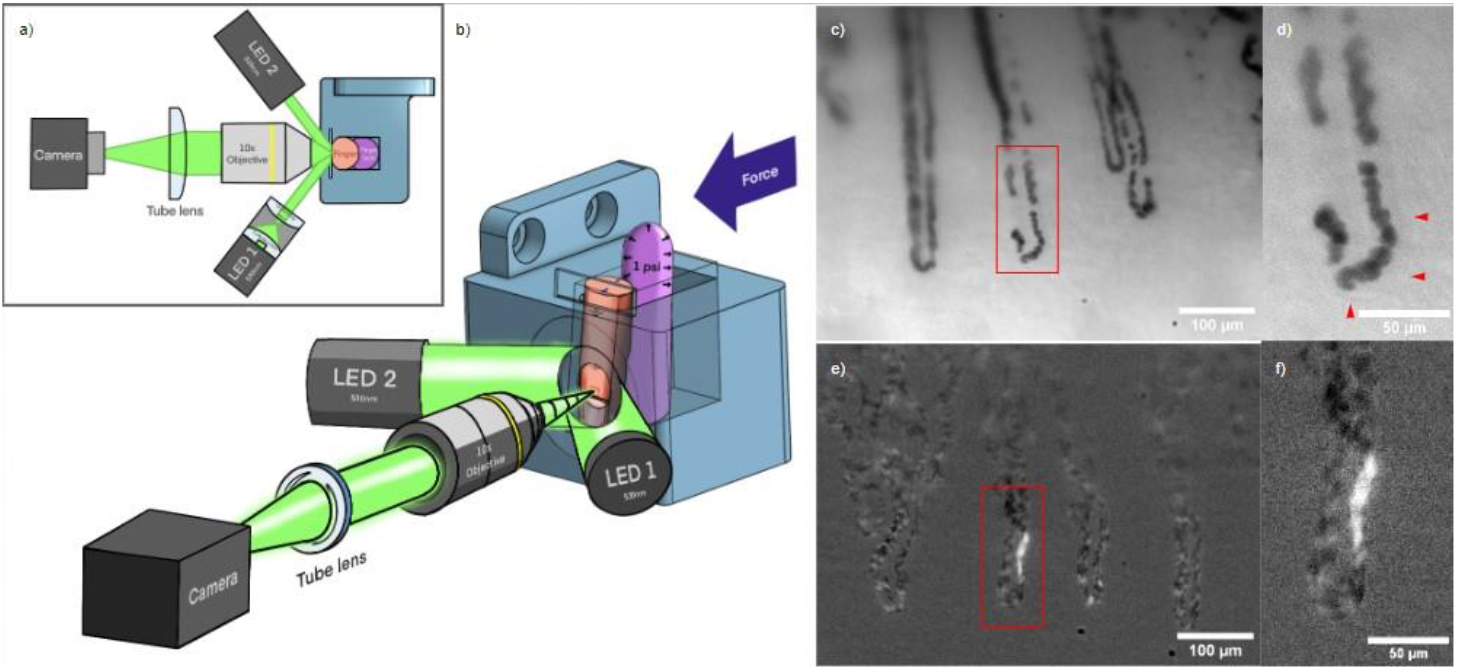
An optical microscope to resolve nailbed capillary blood cells and apply external pressure to the finger: a-b) Top-down diagram and drawing of microscope. Dual 530 nm LEDs illuminate a participant’s finger with collimated light at an oblique angle to minimize reflections. The finger is held against a glass slide with constant force by a balloon (the ’finger lock’) inflated to a specific pressure. This stabilizes the video and controls the flow speed. A 10x infinity corrected objective sends light to a 2” tube lens, which focuses the image on the camera. c) Sample frame showing capillaries at high pressure (1.6 psi) and slow flow. d) Inset showing distinct red-blood cells and faint outline of capillary wall (red triangles). For video, see supplemental information. e) Background-subtracted image showing red-blood cell borders and a white-blood cell gap. f) Inset showing white-blood cell gap.

Participant videos were grouped by location on the nailbed and stabilized, sharpened, and segmented to identify and label individual capillaries (Fig 2 a; Methods). For each capillary, we plotted capillary centerline pixel intensity values with respect to time, creating kymographs in which blood flow is represented by intensity clumps moving diagonally rightward (Fig. 2 b-c); the slopes of the diagonals (Methods) correspond to the average velocity of blood flow.

**Fig. 2.**
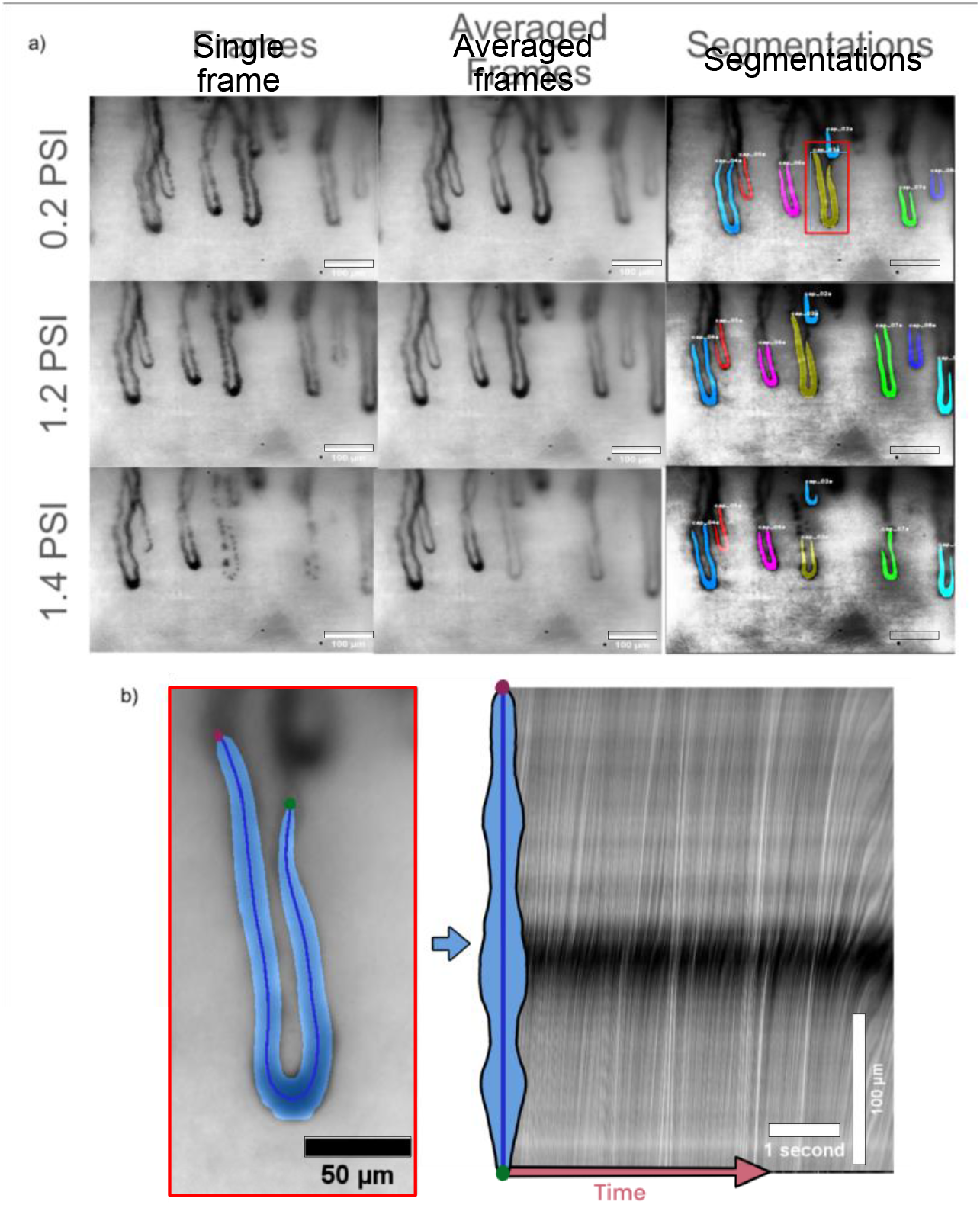
Extraction of capillary blood flow velocity from images: a) Analysis progression at different pressures in the same participant. Registered frames are first averaged by taking the median pixel value of the stack to sharpen capillaries for segmentation. Median images are automatically segmented by hasty.ai before capillaries from different videos at different pressures are matched and labeled. Scale bars are 100 um. b) A centerline is calculated for each segmented capillary. c) The pixels from that centerline are plotted for each frame, making a so-called ’kymograph’ showing blood-packet trajectories over time where the x-axis is time and the y-axis is position on the capillary centerline. The white line on the kymograph shows the calculated average velocity.

### 2.2 Analysis

To compare velocities in the same capillary at increasing and decreasing externally applied pressure, we matched capillaries from the same location across different videos and plotted them with respect to pressure. Because some capillaries evacuate—lose all their blood—or defocus at higher pressures, we selected one capillary per location to serve as the ”representative capillary” for single-capillary tracking. Single-capillary tracking showed differences in participant response with some participants requiring much lower external pressures to slow down and stop blood flow. For example, Participant Eighteen’s blood flow dropped from 1500 um/s to 0 um/s with only 0.4 psi of external pressure (Fig. 3a-d). In contrast, Participant Twenty never had their blood flow stop, even with the full 1.2 psi of external pressure (Fig. 3 i-l). As we released pressure, some participants had their blood flow return symmetrically at the same point that it stopped while others showed an increase in flow rate as it returned, overcorrecting to the restriction in flow. Other capillaries lagged, requiring much lower pressures for blood to return than they needed to stop the flow (Fig. 3 m).

**Fig. 3.**
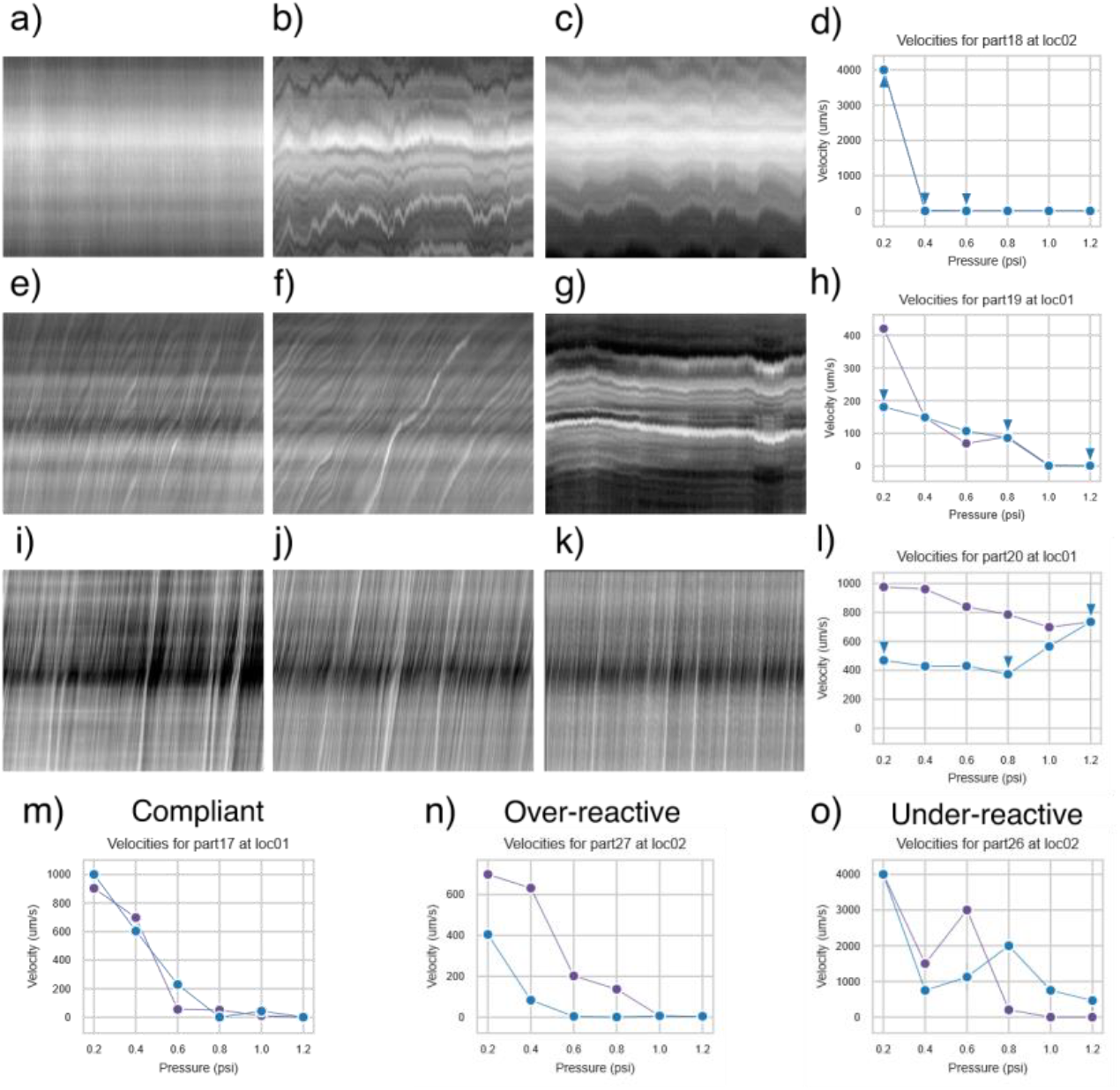
Capillary blood flow at different pressures shows varied responses of participant capillaries to external pressure: a-c) Kymographs from one participant at different pressures, showing very fast blood flow velocity at 0.2 psi before dropping to zero at 0.4 psi; the velocity remained at zero at 0.6 psi. d, h, l, m, n, o) Plots showing average video velocity at the externally applied pressure of that video. In our test, we increased pressure from 0.2 psi to 1.2 psi (videos marked in blue) before decreasing the pressure from 1.2 psi down to 0.2 psi (videos marked in purple). d) Average video velocities at all pressures from the same participant as (a-c). Velocities from (a-c) were plotted with a blue triangle above them because they were measured during the increasing pressure ramp. e,g) Kymographs from a different participant where flow stopped more gradually. h) Average velocities from (e-g) plotted with blue triangles above. i-k) Kymographs from a participant whose blood flow never stops and increases continuously after reaching a minimum value at 0.8psi as pressure is added and then released. This shows reactive hyperemia, where an occlusion causes the body to overreact and vasodilate. l) Average video velocities during the pressure ramp from(i-k) plotted with blue triangle above. m-o) Average video velocity plots showing symmetric (m), overresponsive/hyperemic (n), and laggy capillary responses (o) from different participants.

Given this variation among individuals, we grouped participants by age, blood pressure, and sex and plotted the cumulative density functions (CDFs) of their velocity distributions to see which variables had the biggest effect size. We saw a large separation due to age, with participants under fifty having far more velocities at zero (44%) and a far lower median velocity (80.52 ± 263.32 µm/s) than participants fifty or above (18%, 362.02 ± 288.75 µm/s). Because of this difference in median velocities, we conducted a two-way ANOVA analysis (Table 2) with interactions to analyze the effects of age (grouped as fifty and above and below fifty), systolic blood pressure, and sex on participant median velocity. We found age to be a significant predictor of median velocity (p=0.005), with the fifty and above group showing a 349.6% higher median velocity (362.02 ± 288.751 um/s) than the below fifty group (80.52 ± 263.32 µm/s) (Fig. 4 a). Blood Pressure and sex had smaller (f=1.02, f=2.36) effects than age (f=9.27) and were not significant predictors of median velocity (p=0.321, p=0.136) (Table. 2).

**Table 2.**
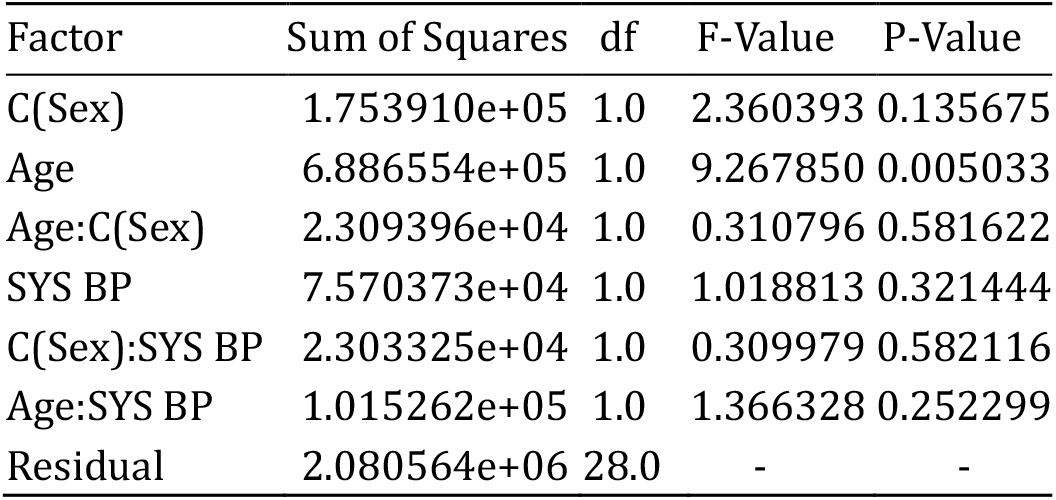
ANOVA results showing the effects of sex, age, and systolic blood pressure on median participant velocity.

| Factor | Sum of Squares | df | F-Value | P-Value |
| --- | --- | --- | --- | --- |
| C(Sex) | 1.753910e+05 | 1.0 | 2.360393 | 0.135675 |
| Age | 6.886554e+05 | 1.0 | 9.267850 | 0.005033 |
| Age:C(Sex) | 2.309396e+04 | 1.0 | 0.310796 | 0.581622 |
| SYS BP | 7.570373e+04 | 1.0 | 1.018813 | 0.321444 |
| C(Sex):SYS BP | 2.303325e+04 | 1.0 | 0.309979 | 0.582116 |
| Age:SYS BP | 1.015262e+05 | 1.0 | 1.366328 | 0.252299 |
| Residual | 2.080564e+06 | 28.0 | - | - |

**Fig. 4.**
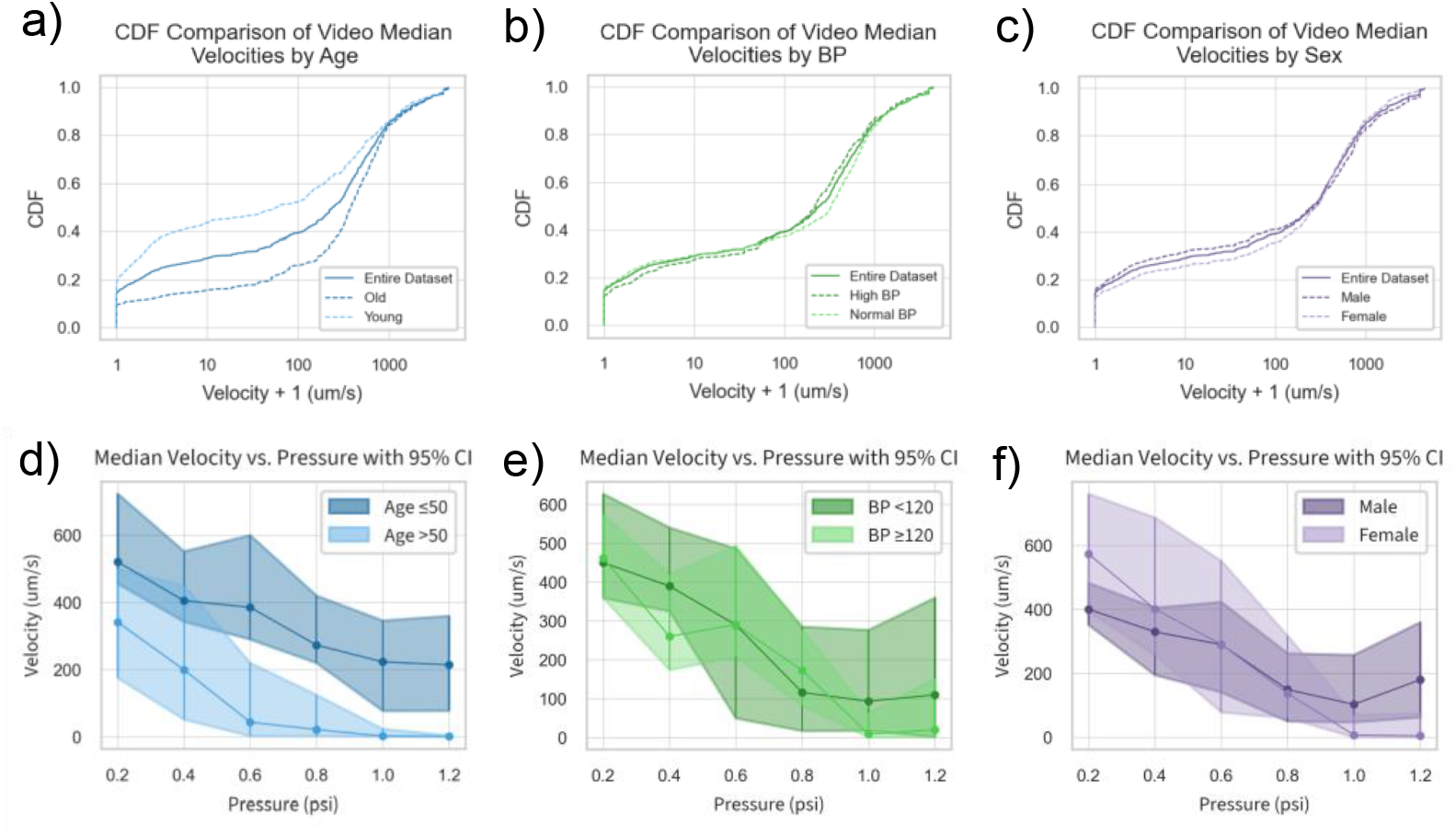
Group blood flow distributions show higher velocities for older participants with mild influence from blood pressure and sex: a) Cumulative Density Function (CDF) plots of age-group blood flow velocity distributions. Participants aged fifty and over had significantly higher median blood flow (ANOVA p-value 0.005). b) CDF plots of blood-pressure-group blood flow velocity distributions. The difference between the low (*<*120 mm Hg) and high (≥120 mm HG) groups was not significant (ANOVA p-value 0.32). c) CDF plots of sex-group blood flow velocity distributions. The difference between sexes was not significant (ANOVA p-value 0.13). d-f) Group 95% median velocity and confidence intervals at each external pressure were calculated by bootstrapping the median of group velocities. P-values for distribution dissimilarity were calculated using the KS Statistic. d) Age groups show large separation in velocity distributions at each external pressure (table 3). e) Blood pressure groups show consistent overlap at each pressure. f) Sex groups show mild separation at low and high external pressures, with women having significantly higher velocities at low pressures and lower velocities at higher pressures.

To evaluate how external pressure affected velocities in each group, we resampled our velocity data at each pressure with replacement to form 1000 bootstrap samples and calculated their median velocities with 95% confidence (Fig. 4). We quantified dissimilarity between group velocity distributions at each external pressure using the two-sample Kolmogorov-Smirnov Test, showing significantly higher velocity distributions at each external pressure for older participants (Table 3). In contrast, we did not see significant differences in velocities between normal and high blood pressure at any external pressure (Fig. 4 d, Table 3). We saw mild separation at low and high external pressures, with women having significantly higher velocities at low external pressures (0.2psi, 0.4psi) and lower velocities at the highest external pressure, 1.2psi (Fig. 4 e, Table 3).

**Table 3.** Kolmogorov-Smirnov (KS) test for velocity distribution dissimilarity at increasing external pressures

| Age |  |  | SYS -BP |  |  | Sex |  |  |
| --- | --- | --- | --- | --- | --- | --- | --- | --- |
| Pressure (psi) | KS Statistic | p-value | Pressure (psi) | KS Statistic | p-value | Pressure (psi) | KS Statistic | p-value |
| 0.2 | 0.329 | 0.00018 | 0.2 | 0.067 | 0.98614 | 0.2 | 0.209 | 0.04623 |
| 0.4 | 0.315 | 0.00056 | 0.4 | 0.150 | 0.31562 | 0.4 | 0.228 | 0.02666 |
| 0.6 | 0.329 | 0.00030 | 0.6 | 0.149 | 0.33106 | 0.6 | 0.126 | 0.51372 |
| 0.8 | 0.329 | 0.00043 | 0.8 | 0.193 | 0.11368 | 0.8 | 0.092 | 0.87087 |
| 1.0 | 0.341 | 0.00020 | 1.0 | 0.215 | 0.05774 | 1.0 | 0.208 | 0.06572 |
| 1.2 | 0.441 | 0.00004 | 1.2 | 0.227 | 0.11361 | 1.2 | 0.267 | 0.03673 |

Because velocities under 1000 um/s best-differentiated age groups, we used a windowed ‘earth-movers distance’ (Methods) to calculate an ‘age-score,’ comparing a participant’s vasculature to the average of the full group (Fig. 5 a). Plotting age-scores with respect to age showed negative age-scores for older participants with four outliers (Fig. 5 b), including two participants with low systolic blood pressures (99, 100). Using age-score as a threshold to classify participants into young (*<*50) and old (≥50) age groups gave a receiver operating characteristic (ROC) with an Area Under the Curve (AUC) of 0.79±0.07 with 95% confidence interval of [0.64, 0.94] (Fig 5 c). Using the median velocity of each participant at 1.2 psi as a threshold gave a ROC with a similar AUC of 0.75± 0.09 with a confidence interval of [0.58, 0.92] (Fig. 5 d).

**Fig. 5.**
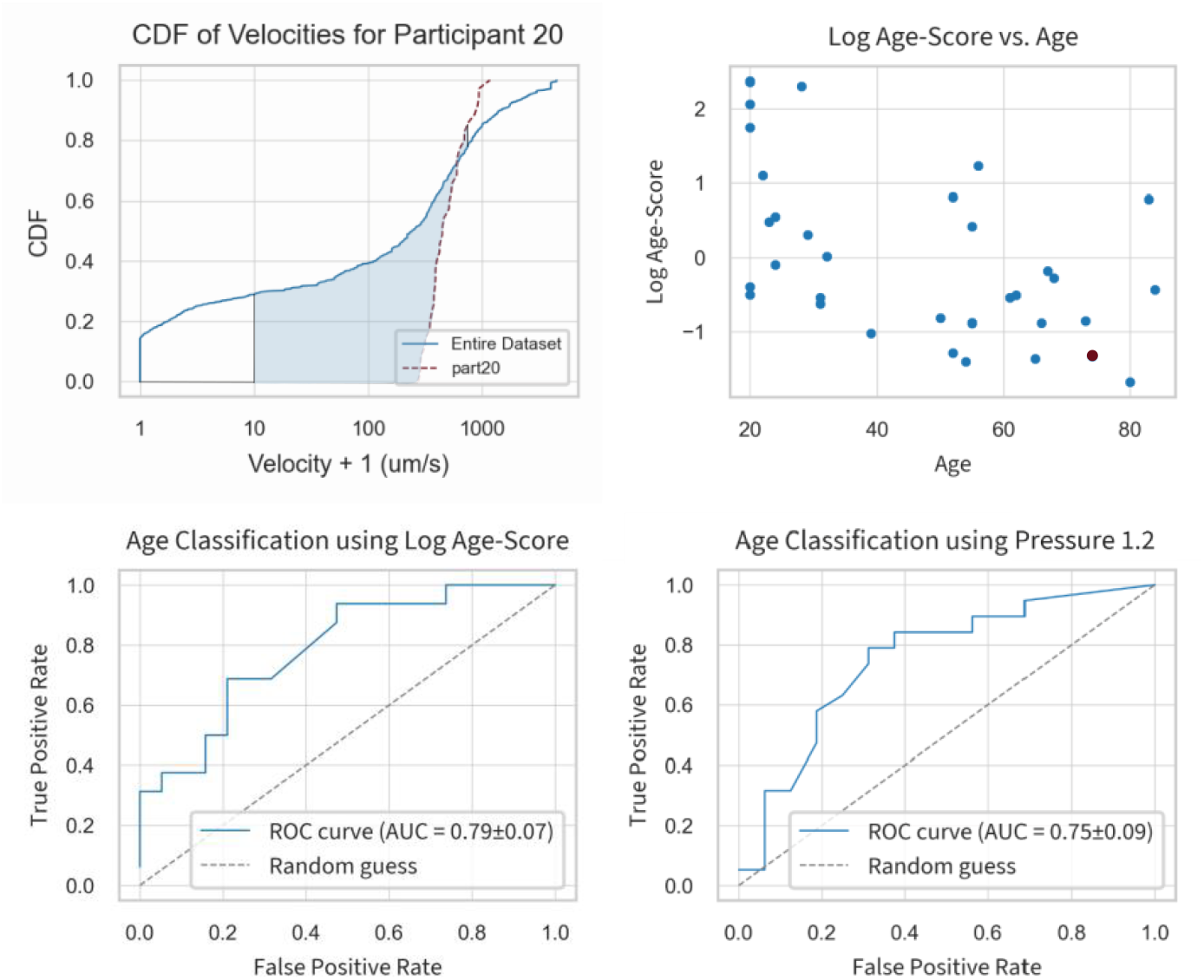
Participant similarity to the population mean, ‘Age-score,’ predicts vascular age: a) ’Age-score’: a windowed (10-750 um/s) earth mover’s distance to compare participant velocity distributions to the average velocity distribution. The participant distribution is in orange and the population distribution is in blue. b) Participant age-scores plotted against age. The participant from part (a) is plotted in red and all other participants in blue. c) ROC curve for Log Age-score. d) ROC curve for Log Median Velocity at 1.2 psi.

## 3 Discussion

Here we demonstrate the ability of our custom-built capillary microscope to systematically slow and measure blood velocities in healthy participants of different ages. Individual participants varied in their response to external pressure, with some participants having blood flow stop and return symmetrically at the same pressure and others showing a hysteresis—either overcorrecting with blood returning too fast or under-correcting with blood returning at a pressure much lower than that which originally stopped flow. An increase in blood flow after occlusion—a reactive hyperemia— is commonly seen in systemic arteries and veins [1, 3, 30] and indicates a responsive but imperfectly elastic vascular system. A slow return of blood flow— impaired reactive hyperemia—suggests endothelial dysfunction, which could result from imbalanced nitric oxide levels or poor vasodilation. We measured a significant increase in the median velocities of participants over 50. This increase in blood flow velocity may stem from previously observed age-related changes in vasculature, such as endothelial cell stiffening[2–4], reduced vascular reactivity [1], or decreasing capillary density [31–34]. Taken together, our instrument is a non-invasive technique to measure age effects in capillaries. Because capillaries show damage and dysfunction before systemic symptoms in various diseases[19–25], we anticipate that this approach will enable early detection of such diseases–motivating future studies in disease diagnosis, monitoring medicine compliance, and continuous tracking of circulatory health.

## 4 Methods

### 4.1 Capillary Microscope

To visualize flow dynamics at 8.418x magnification and 227.8 frames per second (fps) in human nailbed capillaries, we built a custom tabletop microscope using a 10x infinity corrected objective, a 150 mm achromatic tube lens, and a 227.8 fps Basler Camera (Fig. 1a-b). We illuminated the finger with two collimated beams of maximally defocused light from 530 nm green Thorlabs LEDs in the Kohler configuration (Fig. 1a-b). The beams intersected the finger at 30° angles with the nailbed to avoid surface reflections into the camera. We resolved 1951 USAF target lines up to Section 9 subsection 3, giving us a resolution of at least 645.1 lpm (Supp. Fig. 9). To reduce finger shaking, we 3D printed a custom fingerholder chamber and inflated a non-latex glove as a ”finger-lock” to hold the finger against a No. 2 (0.19–0.20 mm thick) glass coverslip with constant pressure, between 0.2 and 2.0 psi.

### 4.2 Pressure Ramp

For each participant, we took videos of nailbed capillaries at 6 different external pressures. Each video was 625 frames, taken at 113.9 fps. After the video was taken, there was a 5-second delay while the camera buffered. During this time, we increased the pressure by 0.2 psi and refocused the camera to record when the camera finished buffering. Hence, we took a 5.49-second video every 10.49 total seconds. The pressure in the inflatable finger-lock was controlled using an Enfield TR-010-v-ex pressure regulator.

### 4.3 Finger Location Selection

We primarily used the ring and pinky fingers for videos because their capillaries are closer to the surface, giving better images. We looked for locations near the cuticle where multiple capillaries were exposed and flowing. Specifically, we used capillaries at the top of the cuticle, where the finger presses on the glass slide. Once we found a flowing capillary, we took a series of videos, choosing one capillary to focus on as we increased the pressure. We adjusted the field of view to keep the chosen capillary in frame. We would select the second location based on the following trends: Ring fingers had larger but deeper capillaries than pinky fingers. If our first location had big capillaries that were too deep, we would check the pinky to see if we could trade size for clarity. If ring finger capillaries were too small, we would try the pointer and then middle fingers for bigger capillaries. Most often, the ring finger was the best choice.

### 4.4 Data Pipeline

Participant videos were grouped by finger location and analyzed to extract velocity data of individual capillaries. To sharpen capillary edges for segmentation, we averaged frames by taking the median of video stacks stabilized using the ”MOCO” ImageJ plugin [35] with a downsample value of 2 (Fig 2 a). We checked each video to ensure the videos were stable and sliced videos if a portion of the video was out of focus or incorrectly stabilized.

### 4.5 Segmentation

We segmented capillaries using a custom-trained ”hasty.ai” [36] neural net, editing segmentations to remove loops and overlapping capillaries (Supplementary Fig 7).

### 4.6 Centerlines

To calculate the centerline of our capillary segmentations, we used the FilFinder skeletonize algorithm [37] and pruned centerlines with a branch threshold of 40 pixels (16.4 um), a minimum capillary length of 50 pixels (20.5 um), and a prune criterion of ‘length’. For the pruning to work correctly, we required segmentations without loops.

### 4.7 Kymographs

To make kymographs, we averaged centerline values using a 4-pixel-radius (1.64 um) circular kernel (Supplementary Fig 6) and plotted them with respect to time. We removed horizontal banding from an image by subtracting a smoothed version of the row means from the image. We obtained lines representing blood packet trajectories from kymograph images by applying a sigma 2 Gaussian blur to the image, using Canny edge detection, and applying the Hough Transform to find lines. We calculated velocities by measuring the average of the slopes of these lines, weighted by the length of the line.

We validated velocities using a custom-made graphical user interface (GUI), which plots the called velocity on top of the kymograph (Supplement Fig 8). We would then manually determine if the velocity was: Correct (T/F), zero/horizontal (T/F), ‘too slow’, or ‘too fast.; If the velocity call was correct, we retained the called value. If incorrect but the kymograph showed horizontal lines (zero flow), we marked the ”Corrected Velocity” as zero. Otherwise, we classified the velocity as ”too fast” or ”too slow.” We then manually annotated these ‘too fast’ and ‘too slow’ velocities by overlaying the following velocities– 20, 35, 50, 75, 110, 160, 220, 290, 360, 420, 500, 600, 750, 1000, 1500, 2000, 3000, 4000 um/s–onto the image and choosing the correct slope.

### 4.8 Median Velocities

To normalize for unequal numbers of capillaries in each video when comparing participants, we calculated the median velocity across distinct capillaries for each video. The median of these medians is the median participant velocity. We used medians instead of means because high-velocity values had a larger uncertainty than smaller values.

### 4.9 Cumulative Density Functions

Cumulative density functions (CDFs) were made using the participant’s median video velocity (seen above). The farther the distribution line is shifted to the right, the faster the distribution of velocities.

### 4.10 Windowed Earth Mover’s Distance

The earth mover’s distance (EMD) is a measure of dissimilarity between distributions defined as the difference in area of two cumulative distribution function (CDF) curves. We used a windowed EMD to measure the difference in area under the curve for the velocities from 10 um/s to 750 um/s. We chose this because velocities higher than 750 um/s had higher uncertainty but could be binned together into a “higher than 750 um/s” bucket. Similarly, values at or below 10 um/s had very similar kymographs and made up a large percentage of velocities. We treated them as the same in this method.

## Data Availability

Data for this manuscript is protected by HIPAA and therefore is unavailable without the consent of the participants.

## Ethical approval

1. Approval: All experimental methods were approved by Stanford institutional review board (IRB) 6 in protocol number 64537.
2. Accordance: All methods were carried out as written in the IRB protocol 64537.
3. Informed consent: Informed consent was obtained from all participants.

## Code availability

All code is available at: https://github.com/gt8mar/capillary-flow

## Author contribution

M.L.F and S.R.Q. conceptualized the study. M.L.F. and S.R.Q. designed the study. M.L.F., S.R.Q., and D.N.C. wrote the IRB protocol. G.R. wrote the capillary-naming pipeline. M.L.F. wrote all other parts of the data processing and analysis pipeline. M.L.F, G.R, J.H.L, S.R.Q., and D.R.C recruited all participants. M.L.F., G.R., and J.H.L. collected, processed, and performed quality control on all data. M.L.F. performed all analyses. M.L.F. and S.R.Q. wrote the manuscript. All authors revised the manuscript and approved it for publication.

### Corresponding author

Correspondence to Stephen R. Quake.

## Supplementary information

## Acknowledgements

We thank G.E. Marti for optics and statistics discussions. We thank J. Lee, T. Chen and S. Cofer for helpful discussions. We thank Hongquan Li for optics and software discussions. We thank K. Medill for electrical circuit input, feedback, and helpful discussions. We thank N. Kozak and A. Marsden for fluid-flow discussions. Funding: This work is supported by the Chan Zuckerberg Biohub. M.L.F. is supported by a National Science Foundation Graduate Research Fellowship, and the Knight-Hennessy Scholars Program.

## Supplementary Figures

**Sup Fig. 1.**
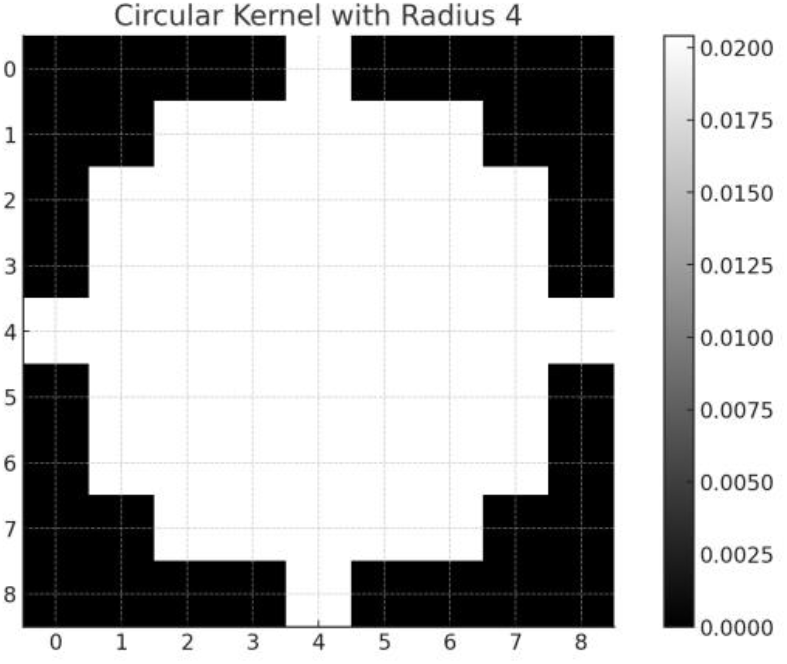
Circle of pixels used to create kymographs.

**Sup Fig. 2.**
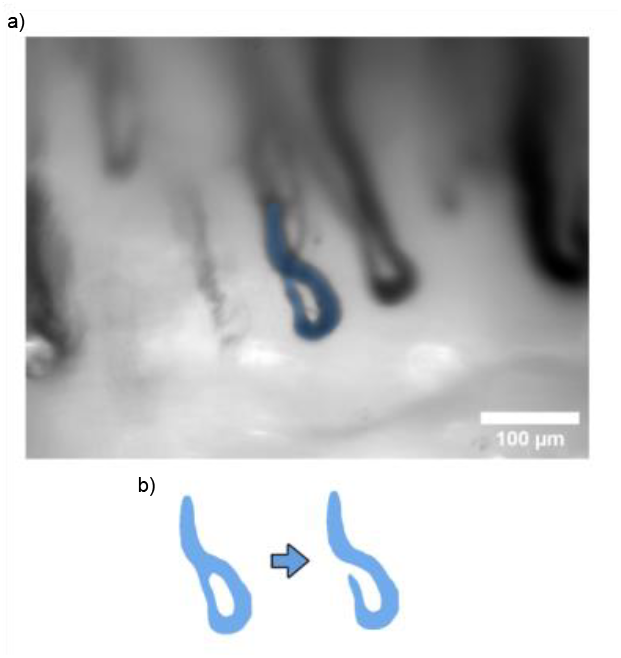
Manual cleaning process to remove loops from capillary segmentations. Our analysis cannot currently prune skeletons with loops.

**Sup Fig. 3.**
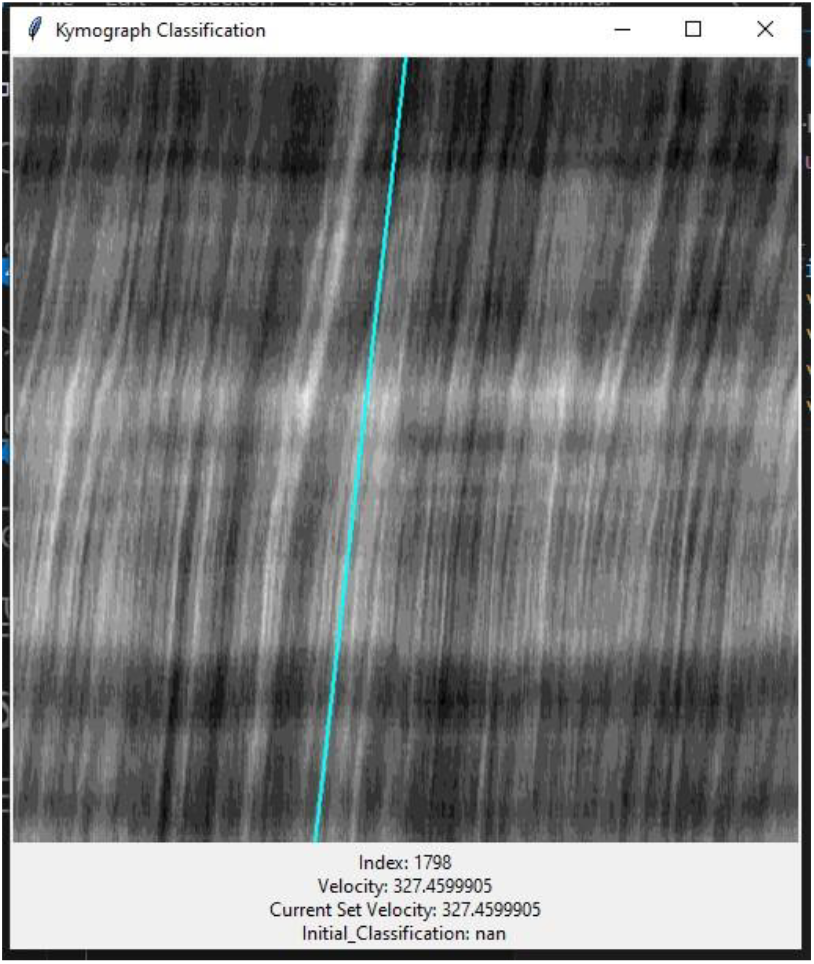
The GUI we use to manually check and call velocities.

**Sup Fig. 4.**
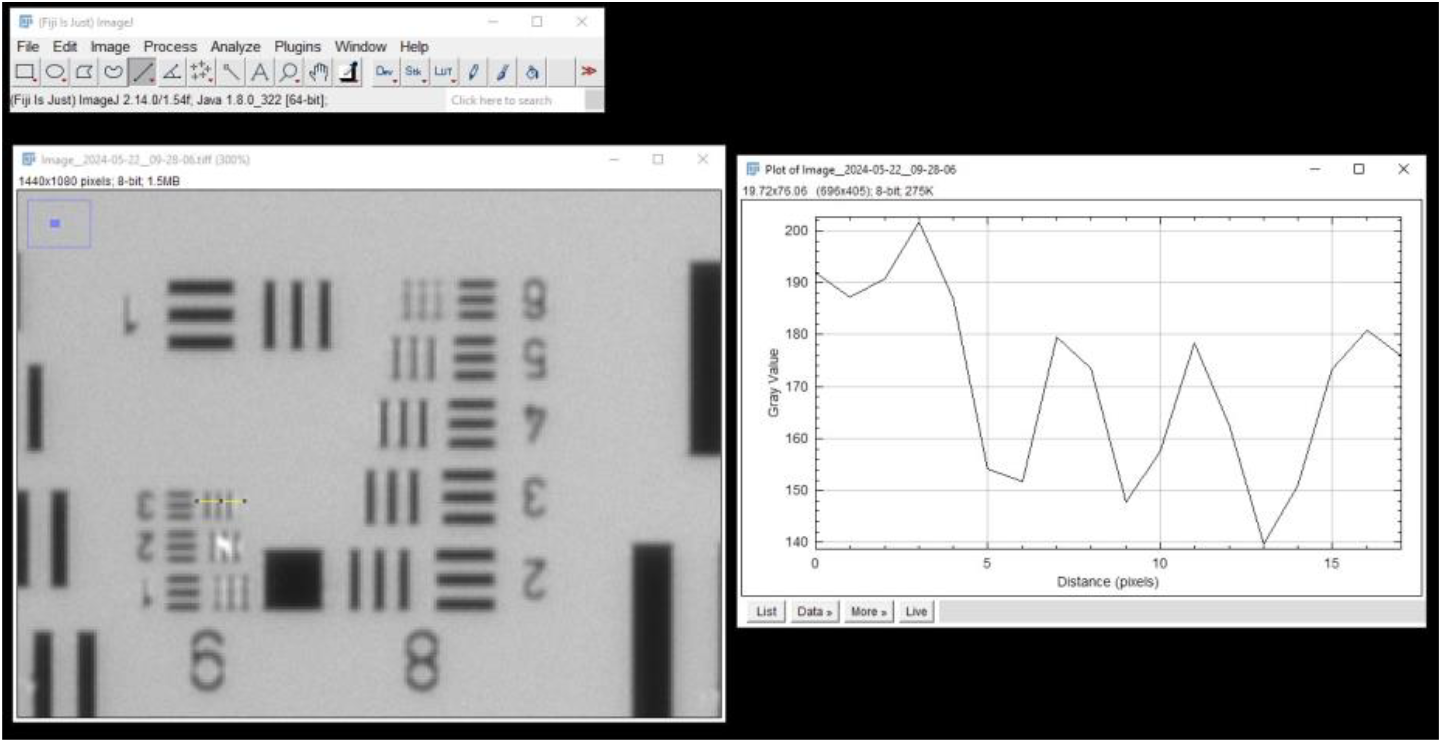
USAF Resolution Target shows resolution exceeds 645.1 lpm

